# Artificial Intelligence in Neuro-Oncology: Assessing ChatGPT’s Accuracy in MRI Interpretation and Treatment Advice

**DOI:** 10.1101/2025.04.22.25326204

**Authors:** Abdullah H. Ishaque, Alexandre Boutet, Shivaprakash B. Hiremath, Matthew P. Mullarkey, Maria Peris-Celda, Gelareh Zadeh

**Affiliations:** Division of Neurosurgery, Department of Surgery, Temerty Faculty of Medicine, University of Toronto, Toronto, Ontario, Canada; Division of Neurosurgery, Krembil Brain Institute, University Health Network, Toronto, Ontario, Canada; Division of Neuroradiology, Joint Department of Medical Imaging, University Health Network, Toronto, Ontario, Canada; Department of Medical Imaging, Temerty Faculty of Medicine, University of Toronto, Toronto, Ontario, Canada; Department of Neurosurgery, Mayo Clinic, Rochester, Minnesota, USA; Princess Margaret Cancer Centre, University Health Network, Toronto, Ontario, Canada

## Abstract

**Purpose:** Large language models (LLMs) have demonstrated advanced capabilities in interpreting text and visual inputs. Their potential to transform oncological practice is significant, but their accuracy and reliability in interpreting medical imaging and offering management suggestions remain underexplored. This study aimed to evaluate the performance of ChatGPT in interpreting T1-weighted contrast-enhanced MRI images of meningiomas and glioblastomas and providing treatment recommendations based on simulated patient inquiries.

**Methods:** This observational cohort study utilized publicly available MRI datasets. Thirty cases of meningiomas and glioblastomas were randomly selected, yielding 90 images (three orthogonal planes per case). ChatGPT-4o was tasked with interpreting these images and responding to six standardized patient-simulated questions. Two neuroradiologists and neurosurgeons assessed ChatGPT’s performance using five-point Likert scales and their inter-rater agreement was evaluated.

**Results:** ChatGPT identified MRI sequences with 91.7% accuracy and localized tumors correctly in 66.7% of cases. Tumor size was qualitatively described in 85% of cases, and the median acceptability was rated as 4.0 (IQR 4.0–5.0) by neuroradiologists. ChatGPT included meningioma in the differential diagnosis for 73.3% of meningioma cases and glioma in 83.3% of glioblastoma cases. Inter-rater agreement among neuroradiologists ranged from moderate to good (κ = 0.45–0.72). While surgical treatment was suggested in all symptomatic cases, neurosurgeon acceptability ratings varied, with poor inter-rater reliability.

**Conclusions:** ChatGPT demonstrates potential in interpreting neuro-oncological MRI images and offering preliminary management recommendations. However, errors in tumor localization and variability in recommendation acceptability underscore the need for physician oversight and further refinement of LLMs before clinical integration.

## Introduction

The use of artificial intelligence (AI) in oncology research has expanded rapidly in recent years. Publicly available conversational chatbots powered by large language models (LLMs), such as ChatGPT and Claude AI, have been evaluated as tools for cancer diagnosis,^1^ patient education,^2^ and structuring oncological data.^3^ As these technologies are integrated into healthcare delivery, AI—particularly LLMs—is anticipated to have a profound impact on oncology and broader medical practice.

Studies assessing chatbot diagnostic performance have typically relied on publicly available clinical data as input. A recent study demonstrated that ChatGPT achieved 87% accuracy in addressing weekly “Image Challenge” cases from the *New England Journal of Medicine*.^4^ Similar evaluations have incorporated imaging findings from radiology reports,^5,6^ pediatric case challenges,^7^ and ophthalmic images.^8^ Over the past year, several studies have also examined the quality of treatment recommendations provided by chatbots for oncologic conditions. While chatbots show a degree of understanding regarding treatment guidelines, they currently lack the reliability needed for treatment recommendations.^9-11^ Nevertheless, incremental improvements in newer ChatGPT versions suggest that LLMs could soon play a role in the management oncology patients.^12^

Most prior research has relied on textual inputs to evaluate chatbots, given technical limitations. The latest version of ChatGPT (ChatGPT-4o) now accepts both text and visual inputs, allowing for the interpretation of images, not solely radiology reports. This functionality has been tested with chest X-rays^13^, CT scans,^14^ mammograms,^15^ and MRI for detecting stroke,^16^ albeit with restrictions two-dimensional (2D) inputs. In neuro-oncology, only one study has evaluated ChatGPT’s accuracy with CT and MRI images.^17^ Authors of that study used challenging cases published in *Clinical Neuroradiology* as input data, which included patient symptoms, CT and MRI images, and radiology reports, to evaluate the diagnostic performance of ChatGPT against radiologists. The results demonstrated that including images as input yielded a higher diagnostic performance compared to text alone, but overall, its accuracy was lower than radiologists’ assessments. In particular, radiologists were better at providing differential diagnoses.

As patients gain increased access to their personal health records, including medical images, it is likely they will turn to AI-based tools for medical opinions before consulting physicians. Recent surveys indicate a growing public inclination to use these technologies for personal health inquiries.^18^ This trend is especially relevant in oncology, where patients frequently seek second opinions that can impact management and outcomes.^19^ Therefore in this study, we sought to evaluate ChatGPT’s performance in (1) interpreting MRI images of meningiomas and glioblastomas, which are the most common primary central nervous system tumours, and (2) offering treatment recommendations.

## Methods

This study did not require research ethics board approval as it utilized publicly available data. All MRI data were obtained from the Brain Tumour Segmentation (BraTS) challenge that aims to develop MRI segmentation tools specifically for intracranial tumours.^20-22^ BraTS challenges are supported by MedPerf, an open platform for benchmarking AI models in medicine.^23^ The study was designed and reported following the Strengthening the Reporting of Observational Studies in Epidemiology (STROBE) guidelines for cohort studies.^24^

MRI data were sourced from the BraTS 2023 Meningioma and Adult Glioma challenges.^20,22,25^ These datasets were acquired from multiple centers across North America with varying acquisition parameters and data quality. Thirty cases of meningiomas and thirty cases of glioblastomas were randomly selected from the training datasets of their respective challenges. For each case, a two-dimensional (2D) JPG image of the post-contrast T1-weighted MRI sequence was captured from each orthogonal plane to represent the tumour’s maximum diameter (Figure 1). The image resolution for all cases was at least 1 x 1 x 1 mm. Specific acquisition parameters were not available as part of the dataset. The final dataset comprised 90 images (60 tumours × 3 orthogonal planes), which were used as input.

**Figure 1:**
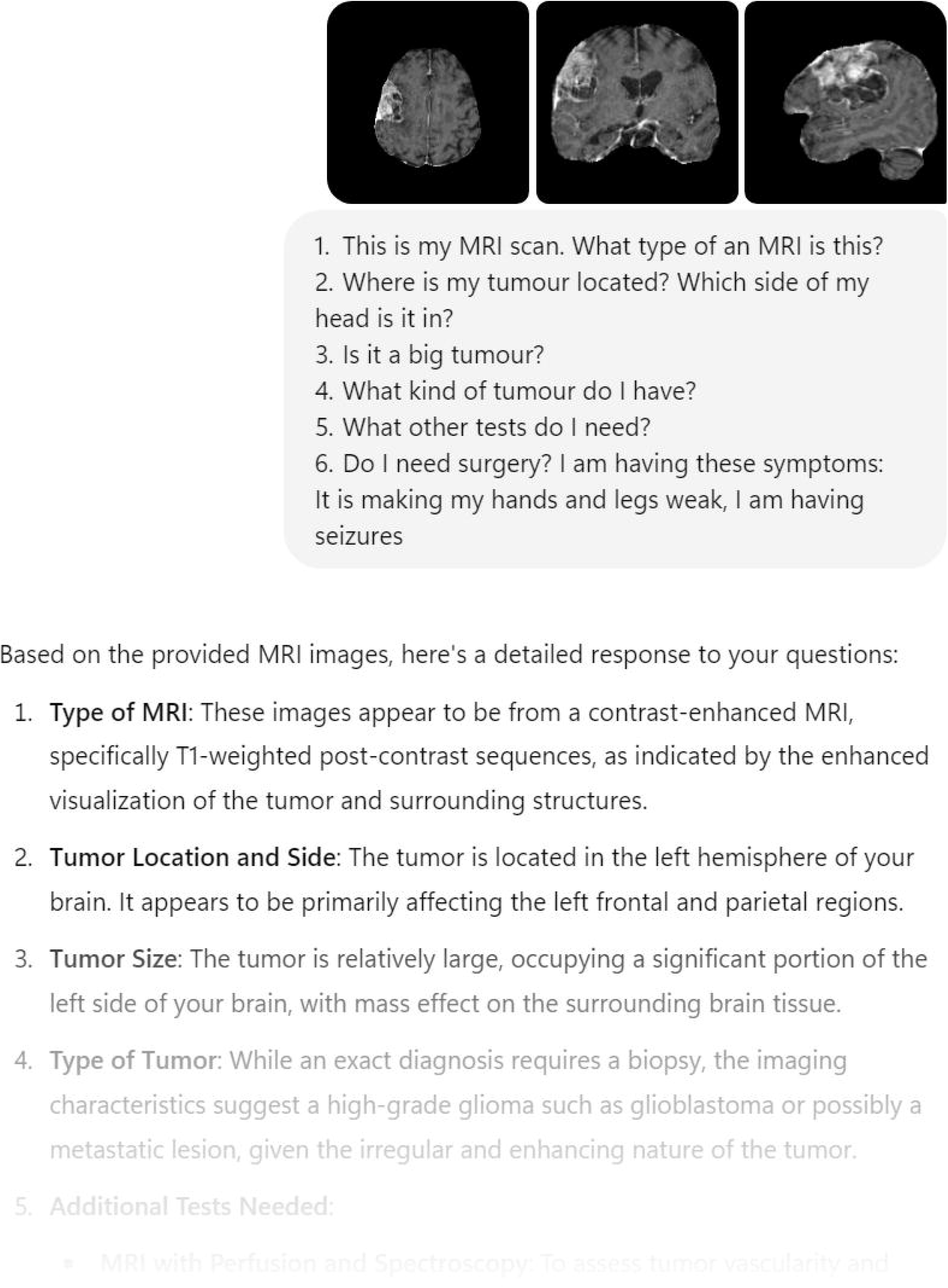
Example of input images. Sample meningioma (A) and glioblastoma (b) cases shown with axial, coronal, and sagittal T1-weighted contrast images in radiological convention (from left to right).

Six input questions were developed to simulate patient questions:

1. What type of MRI is this?
2. Where is my tumour located? Which side of my head is it on?
3. Is it a large tumour?
4. What type of tumour do I have?
5. What other tests do I need?
6. Do I need surgery?

The last question was paired with symptoms attributable to the tumour’s location and size: no symptoms, headaches and nausea, weakness in limbs, seizures, difficulty speaking, and visual impairment (Figure 2).

**Figure 2:**
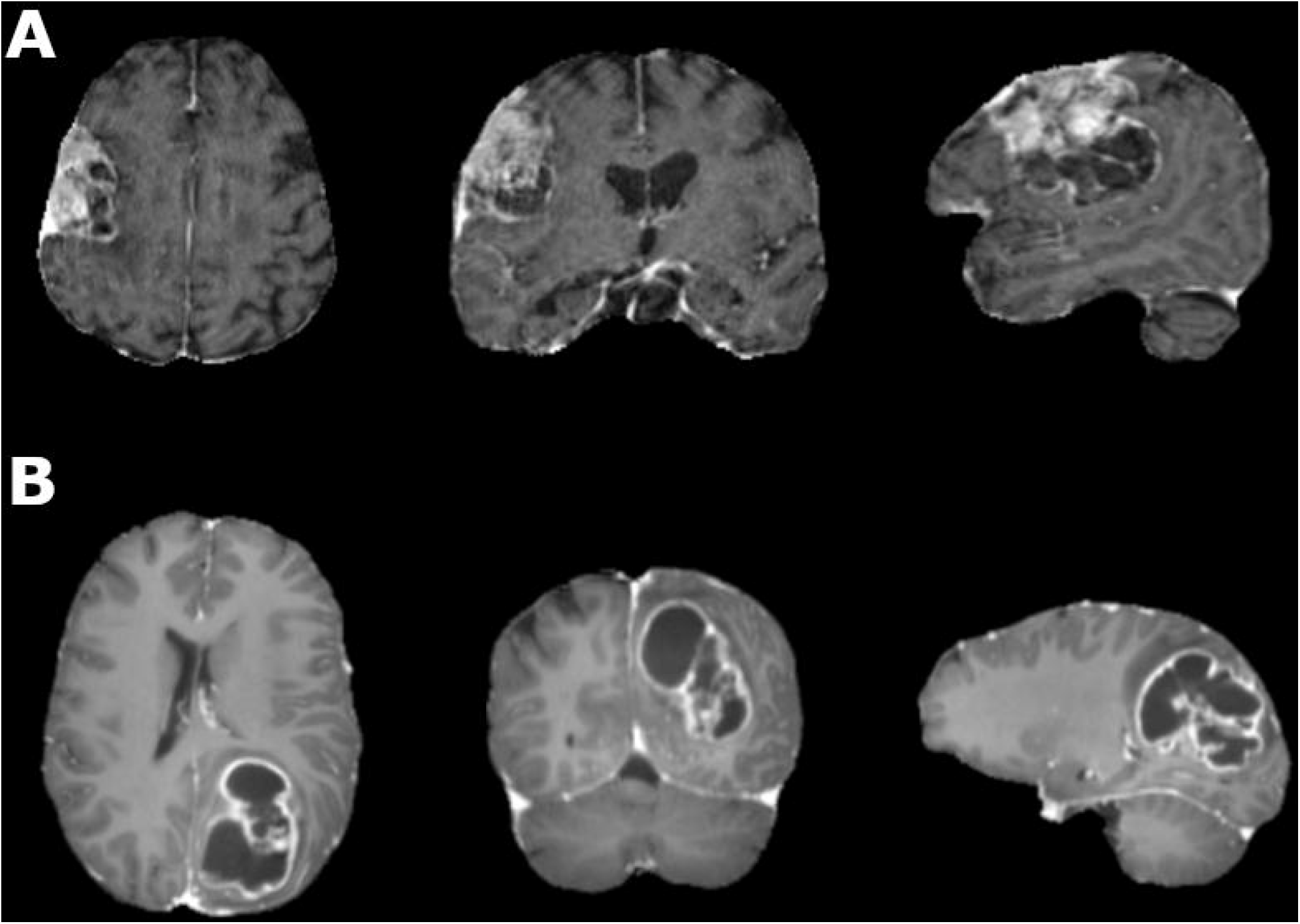
Sample responses from ChatGPT following input data.

For each case, the combined data—three 2D images, input questions, and associated symptoms—were provided to ChatGPT-4o. This task was completed between September and October 2024 using a single personal computer. A new ChatGPT-4o instance was launched for each case, totaling 60 separate instances. No additional prompts, clarifications, or questions were provided beyond the initial input for data analysis.

Statistical analysis was conducted using MedCalc 23.1.5. Two board-certified neuroradiologists assessed ChatGPT’s responses for tumour location, size, and diagnosis. Tumour location was evaluated in a binary manner. The acceptability of the response to tumour size and diagnosis was evaluated on a five-point Likert scale ranging from 1 (strongly disagree) to 5 (strongly agree) by two board-certified neuroradiologists (AB and SBH). ChatGPT’s recommendations for investigations and surgical intervention were evaluated by two board-certified neurosurgeons (MPM and MPC) on a five-point Likert scale ranging from 1 (strongly disagree) to 5 (strongly agree). Inter-rater agreement was evaluated using Cohen’s weighted kappa (κ). Mann-Whitney tests were used to assess for differences between assessors’ ratings with statistical significance set at *p* < 0.05.

## Results

In the 30 meningioma cases, 26 were supratentorial, and four were in the posterior fossa. Among glioblastoma cases, 10 were in the frontal lobe, 13 in the temporal lobe, six in the parietal and occipital lobes, and one in the posterior fossa. ChatGPT identified the input images as post-contrast T1-weighted MRI sequences in 91.7% (55/60) of all cases. Other responses included T1-weighted MRI without contrast, diffusion-weighted MRI, or CT scans. ChatGPT accurately identified the lobe of the brain where the tumour was located in 60% (18/30) of meningiomas and 73.3% (22/30) of glioblastomas. However, it correctly identified the tumour’s side in only 40% (12/30) of meningiomas and 30% (9/30) of glioblastomas.

ChatGPT provided qualitative size descriptions in 85% (51/60) of all cases: 76.5% (39) labeled as “large,” 15.7% (8) as “medium” or “moderate,” and 5.9% (3) as “small.” In remaining cases, tumours were described as “significant” or “sizable.” The median acceptability of ChatGPT’s responses to tumour size was graded as 4.0 by both neuroradiologists, with interquartile ranges of 4.0 – 5.0 and lowest rating of 2. The strength of inter-rater agreement was moderate with κ = 0.45 [95% confidence interval (CI) = 0.24 – 0.65, standard error (SE) = 0.10]. There was no statistically significant difference between the raters’ assessments.

ChatGPT suggested a meningioma diagnosis in 73.3% (22/30) of meningioma cases: in three cases, it provided a single meningioma diagnosis, while in 27 cases, it offered multiple differential diagnoses. The misdiagnosed tumours were in the posterior fossa, anterior skull base, and temporal region. Incorrect differentials for posterior fossa meningiomas included hemangioblastoma, ependymoma, vestibular schwannoma, medulloblastoma, or pilocytic astrocytoma; for anterior skull base meningiomas, pituitary adenomas or craniopharyngiomas; and for temporal meningiomas, glioblastomas or metastases. For glioblastoma cases, ChatGPT included glioma in the differential diagnosis in 83.3% (25/30). Incorrect diagnoses involved meningiomas, metastases, low-grade gliomas, craniopharyngiomas, and pituitary adenomas. The median acceptability of ChatGPT’s responses to diagnosis was graded as 4.0 by both neuroradiologists, with interquartile ranges of 4.0 – 5.0 and the lowest rating of 1. The strength of inter-rater agreement was good with κ = 0.72 (95% CI = 0.56 – 0.88, SE = 0.08). There was no significant difference between the raters’ assessments.

In nearly all cases (96.7%, 58/60), ChatGPT recommended advanced MRI sequences for tumour evaluation. Most common were MR spectroscopy (63.8%, 37/58) and functional MRI (34.5%, 20/58). Other suggested tests included perfusion MRI, CT scans, blood tests, and tumour biopsies. The median acceptability of ChatGPT’s responses to investigations was graded as 4.0 by MPM (interquartile range = 4.0 – 5.0, lowest rating = 2, and highest rating = 5) and as 3.0 by MPC (lowest rating = 2, and highest rating = 5). There was a significant difference between the raters’ assessments (*p* < 0.001). The strength of inter-rater agreement was poor with κ = 0.04 (95% CI = -0.04 – 0.12, SE = 0.04).

For 17 meningioma cases paired with symptoms and all glioblastoma cases, ChatGPT recommended surgery to alleviate symptoms and obtain a tissue diagnosis. In asymptomatic cases, a conservative approach was suggested; in three instances, surgery was additionally advised if tumour growth or neurological sequelae were anticipated with continued observation. For both tumour types, ChatGPT recommended consultations with a neurosurgeon, or neuro-oncologist for a comprehensive review. MPM rated strongly agreed (5 rating) with ChatGPT’s responses to treatment recommendation in all 60 cases. MPC, on the other hand, rated the acceptability of ChatGPT’s responses to treatment recommendation with a median of 4.0 (interquartile range = 4.0 – 5.0, lowest rating = 3, and highest rating = 5). There was no inter-rater agreement for treatment recommendations because MPM strongly agreed with ChatGPT in all instances. Consequentially, there was also a significant difference between the raters’ assessment (*p* < 0.001).

## Discussion

This study assessed the capability of publicly available, untrained ChatGPT in interpreting brain tumour MRI and providing basic medical opinions. We aimed to assess its potential as a future point-of-care medical imaging interpretation tool for brain tumours. Our findings indicate that the LLMs powering ChatGPT are not yet capable of independently interpreting images or providing reliable medical opinions for common brain tumors. We found that ChatGPT correctly identified post-contrast T1-weighted sequences from 2D MRI images with 92% accuracy, despite not being able to process videos or complete imaging datasets in DICOM or NIfTI formats, which are typically used in neuroimaging research. Within the limitations of 2D orthogonal images, ChatGPT demonstrated 66.7% accuracy in identifying tumour locations. Studying multiple brain lesions with chatbots, such as in brain metastases, is challenging due to the limitation of 2D visual input. Deep learning algorithms trained on large datasets have achieved over 95% accuracy in identifying and labeling heterogeneous MRI data.^26,27^ Additionally, current state of the art deep learning models are able to localize and segment brain tumours such as glioblastomas,^28^ meningiomas,^29,30^ and metastases with over 90% accuracy.^31,32^ It is plausible that a trained version of GPT could improve the accuracy of MRI sequence labeling and tumour localization. Interestingly, ChatGPT’s performance in identifying the correct side of the brain for tumour location was notably poor. In a post-hoc query, ChatGPT responded that it uses radiological convention, yet this task—considered relatively straightforward—was poorly executed. This may be due to “artificial hallucination,” where the model generates outputs unrelated to the input.^33^ We hypothesize that ChatGPT may not consistently follow its rules and could benefit from explicit instructions at the start of each instance to prevent confusion. This remains to be examined in future studies.

In cases where meningiomas were incorrectly diagnosed, ChatGPT provided a general differential for posterior fossa tumours, or typical lesions for anterior skull base, or sellar regions. Similarly, in the temporal region, tumours were often misdiagnosed as glioblastomas or metastases. ChatGPT’s diagnostic process, therefore, appears to involve first localizing the tumour and then generating a differential based on location. The outputs are dependent on the input questions, which in this case were from a patient seeking medical advice. In separate instances, we asked ChatGPT to provide the “most likely diagnosis” for these images, and it was able to diagnose meningiomas and glioblastomas in cases with high-resolution MRI scans.

Previous studies evaluating ChatGPT’s diagnostic accuracy have relied on patient vignettes including clinical history, physical exam, and radiology reports published in journals.^5,6,17^ ChatGPT’s diagnostic accuracy is known to increase when information from reported image findings is paired with patient history and physical exam.^6^ A recent study using MRI images and patient history found that ChatGPT performed worse than radiologists in providing differential diagnoses for intracranial pathologies.^17^ Notably, their study used patient history vignettes, while ours focused on specific symptoms linked to input images. Additionally, our study is focused on 60 cases of commonly presenting meningiomas and glioblastomas, whereas the previous study evaluated a heterogenous set of challenging pathologies among 32 cases.

The current guidelines recommend surgical resection for symptomatic or enlarging meningiomas.^34^ For glioblastomas, the guidelines recommend surgical resection followed by chemotherapy and radiation.^35^ From a public point of view, ChatGPT provided suggestions for surgical management, both in asymptomatic and symptomatic tumours. There was a substantial difference in how neurosurgeons evaluated ChatGPT’s responses, highlighting the complexity of managing neuro-oncology patients. Studies have shown that neuro-oncology management patterns vary significantly across institutions, particularly in decisions regarding treatment strategies for elderly neuro-oncology patients,^36,37^ and surgical resection, the extent of resection, or whether to perform a biopsy.^38-40^ These variations are reflected in our findings, which demonstrate that ChatGPT, in its untrained form, is not suited to provide personalized medical interpretation or treatment recommendations. Crucially, in the studied scenarios, ChatGPT did not participate in a patient-centered dialogue to extract additional pieces of information to supplement clinical-decision making such as symptom duration, severity, and functional impact of symptoms. It has to be highlighted that ChatGPT also did not ask clarifying questions about the patient’s age or medical status, factors that are known to impact clinical decision making and outcomes in managing meningioma and glioblastoma patients.^41-43^ With the capability of ChatGPT and other LLMs trained on clinical data and specific parameters, future application of this technology could include customization by neurosurgeons and institutions alike to their preferred practices. This is particularly important as various specialties move towards creating customized LLMs for their own purposes.^44,45^

With the advent of GPT-4o and similar multimodal LLMs, there is an anticipated surge in research exploring the performance of LLMs in interpreting medical images. This is critical to the field of neuro-oncology because imaging is the first investigation performed for tumour diagnosis. While initial reports in chest X-ray^13^ and basic neuroimaging interpretation^17^ show promise, LLMs are not yet ready for clinical use in the field of radiology. Despite their potential, significant limitations must be addressed before these models can be reliably integrated into patient care.

One critical limitation is the risk of misinterpretation. Although AI technologies will likely improve their diagnostic performance in image analysis, the inherent risk of misinterpretation will necessitate physician oversight in real-world scenarios. Such errors highlight that at this early stage of integrating AI into medical care, there are potential dangers of over-reliance without sufficient validation and supervision. Integration of AI in healthcare workflows will therefore necessitate the evolution and development of new roles and guidelines for clinicians. Additionally, there is the potential risk of error propagation and the erosion of clinician expertise. Over-reliance on AI recommendations could lead clinicians to overlook critical clinical cues or second-guess their judgment. If AI-generated interpretations are accepted without critical evaluation, misdiagnoses may become more frequent which may compromise patient safety. A salient concern common to most medical AI applications is data and patient privacy. Risks of data breaches and dissolution of privacy are central to discussions in medical AI and are yet to be effectively resolved. To mitigate these risks, it is crucial to establish protocols that ensure AI outputs are thoroughly reviewed and validated.

Despite limitations, however, LLMs hold promise in specific applications, such as providing second opinions. Second opinions are a crucial aspect of oncologic neuroradiology practices as they significantly impact the accuracy of image interpretation and subsequent patient management.^46,47^ LLMs could potentially streamline this process by offering preliminary assessments or highlighting areas of concern that warrant further review by neuroradiologists. By serving as an adjunct tool rather than a replacement, LLMs may enhance diagnostic confidence and reduce the likelihood of oversight, especially in resource-limited settings or high-volume practices.

### Limitations

This study relied on simulated patient scenarios instead of real patient data for symptoms and questions. In actual use, a patient would likely engage in a more dynamic conversation with ChatGPT, exploring multiple facets of treatment, such as surgical options, chemotherapy, and radiation. Future research could involve trials comparing real patients’ interactions with ChatGPT against their discussions with physicians to better understand the chatbot’s utility in patient consultations. Lastly, the study was focused on T1-weighted post-contrast MRI that served as input for ChatGPT. A significant part of radiological image interpretation involves evaluating multiple sequences. Iin attempt to reflect potential patient use, we opted to include only one imaging sequence that represents the tumour clearly instead of uploading multiple sequences.

This study utilized publicly available data from the BraTS challenge, which omits facial and skull features from MRIs, ensuring patient privacy. Integrating AI in healthcare raises complex ethical and legal considerations that future studies should address comprehensively.

## Conclusion

This cohort study underscores the current state and potential of ChatGPT and similar chatbots in providing preliminary assessments of neuro-oncological MRI images and offering basic management suggestions directly to patients. While current technology remains in its early stages, it is not yet suitable for clinical application. It holds promise that with further advancements including training with large datasets, it is possible to pave the way for AI’s integration into healthcare as an adjunct for patients for point of care image interpretation and medical advice.

## Data Availability

All data produced in the present study are available upon reasonable request to the authors.

## Statements & Declarations

### Authors Contributions

1. Experimental design: Abdullah H. Ishaque, Gelareh Zadeh
2. Implementation and analysis: Abdullah H. Ishaque, Alexandre Boutet, Shivaprakash B. Hiremath, Matthew P. Mullarkey, Maria Peris-Celda
3. Interpretation of data: Abdullah H. Ishaque, Gelareh Zadeh
4. Writing and editing the manuscript: Abdullah H. Ishaque, Alexandre Boutet, Shivaprakash B. Hiremath, Matthew P. Mullarkey, Maria Peris-Celda, Gelareh Zadeh

### Data Availability

The MRI data used for the study is available online as part of the BraTS challenge

### Ethics Approval

Ethics approval was not required for study as the data was obtained from a publicly available dataset, as confirmed by the local ethics board

